# Covariate Adjusted Logit Model (CALM) for Generating Dose-Response Curves from Observational Data with Applications to Vaccine Effectiveness Trials

**DOI:** 10.1101/2025.02.18.24319273

**Authors:** Nong Shang, Stephanie Schrag, Rebecca Kahn, Julia Rhodes

## Abstract

Establishing dose-response relationships from observational data is challenging due to confounding and sample selection bias. Standard causal methods adjust for confounding but typically require knowledge of covariate distributions in the target population—often via a well-defined probability sampling scheme. We propose the Covariate Adjusted Logit Model (CALM), which generalizes log-linear structural mean models for binary exposures to continuous exposures by modeling a relative dose-response curve anchored to a baseline level. By separating this curve from the null disease risk (NDR) at baseline, CALM enables valid inference under biased sampling while adjusting for confounding effects. A Gibbs sampler—the All-or-Nothing algorithm—is introduced to support Bayesian modeling, drawing on a vaccine-effect-inspired interpretation of the relative dose-response curve. Simulation studies demonstrate that CALM recovers dose-response relationships more accurately in the presence of bias and confounding. In vaccine trials, where confounding covariates affect immune responses differently across study arms, CALM provides a more accurate and robust antibody–disease curve to serve as a surrogate for evaluating vaccine effectiveness.

## 1. Introduction

Dose-response curves are widely used in biomedical, environmental, and epidemiological research to characterize the relationship between a continuous exposure variable and a binary or continuous outcome. Beyond predicting individual or population-level risk, dose-response models serve several practical purposes: identifying thresholds associated with clinically meaningful outcomes, extrapolating risk to unobserved exposure levels, and performing scenario analyses to support intervention strategies.

Traditional dose-response models are often based on data from controlled experiments, where exposure levels are pre-specified and environmental factors are either randomized or tightly regulated. Daniels et al. [1] provide a comprehensive overview of commonly used parametric models in such settings. In contrast, observational studies introduce additional complexities. The lack of randomization leaves results vulnerable to confounding, and sampling processes may induce selection bias—particularly when the sample is not representative of the target population in terms of confounding covariate distributions. Examples include studies of sun exposure and skin cancer risk (Kricker et al. [2]), air pollution and mortality (Di et al. [3]), and vitamin D levels and coronary heart disease (Wang et al. [4]; Crowe et al. [5]). Another motivating example arises from our own work on maternal immunization against Group B Streptococcus (GBS). To establish antibody-based correlates of protection for GBS vaccines, multinational studies have collected observational data on infants’ antibody levels (exposure), infant GBS disease status (outcome), and maternal and infant covariates (Gilbert et al. [6]; Izu et al. [7]; Madhi et al. [8]; Rhodes et al. [9]).

A common strategy to address confounding in observational studies is to adopt a counterfactual framework, where the estimand of interest is the average counterfactual outcome (*Y*) at dose level *E*[*Y*(*d*)]. With presence of observed confounder *X*, and under assumptions of no unmeasured confounding and conditional exchangeability, *E*[*Y*(*d*)] can be estimated either by marginalizing over *X* (i.e. g-formula) or through generalized propensity score methods for continuous exposures (Imbens [13], Hirano and Imbens [14], Brown et al. [15], Fong et al. [16]). However, both strategies rely on accurate knowledge of the marginal distribution of the confounders—either explicitly, in the g-formula, or implicitly in the weighting used by propensity score methods. If the marginal distribution of *X* is unknown or distorted—due to sample selection bias, for example—these standard approaches can fail.

One way to address both confounding and sample selection bias is to constrain the definition of confounding through a modeling framework that avoids reliance on the marginal distribution. A well-known example for binary exposures is the log-linear structural mean model (Robins [14]; Dukes and Vansteelandt [15]; Robins[16]). This approach has two features: it targets the relative effect rather than the absolute risk, and it models the relative effect so that the conditional causal relative risk (given *X*) equals the marginal causal relative risk, assuming conditional exchangeability.

In this paper, we extend this idea to continuous exposure exposures by developing a modeling framework that defines a relative dose-response function with respect to a baseline dose level. Analogous to the relative risk ratio in the model, we define a relative dose-response function with respect to a baseline dose level. We refer to this generalization as the Covariate-Adjusted Logit Model (CALM). While we term “logit” reflects historical usage of logit functions in dose-response modeling, CALM is a flexible framework that accommodates both parametric and non-parametric specifications.

Section 2 introduces the model formally, along with its interpretation in the context of vaccine effectiveness research. In particular, we show that the relative dose-response curve can be interpreted as the proportion of individuals remaining susceptible at each dose level, assuming full susceptibility at the baseline dose level. This interpretation motivates a novel Gibbs sampling algorithm—the All-or-Nothing (AoN) algorithm—which enables more complex modeling of the relative dose response curves.

Section 3 presents simulation studies that evaluate CALM’s ability to recover the relative dose-response curve under various degrees of confounding and sample selection bias. These simulations are followed by an application to real data from an interim analysis of our maternal GBS vaccine study.

Section 4 further explores CALM’s application in vaccine clinical trials, where immunologic surrogates are often used in place of clinical endpoints. Through extending the simulation design in Section 3 to simulate a vaccine with a given vaccine effectiveness (VE), we evaluate the performance of CALM-based curves in estimating vaccine effectiveness and compare them with other commonly suggested approaches. Our results show that CALM consistently provides more accurate and robust VE estimates, particularly in the presence of sample bias or confounding.

Finally, Section 5 concludes with a discussion of CALM’s methodological contributions and outlines directions for future research and application.

## 2. Covariate Adjusted Logit Model (CALM)

### 2.1. Definitions and interpretations

Rather than modeling the absolute dose-response estimand *E*[*Y*(*d*)], we consider a relative dose-response relationship 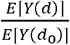, where *d*_0_ is the baseline exposure level. When only two exposure levels exist, 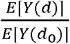 represents the causal risk ratio. In the presence of a confounding covariate *X*, a standard approach is to model the conditional causal risk ratio 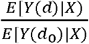 using a log-linear structural mean model (Robins [14]; Dukes and Vansteelandt [15]):

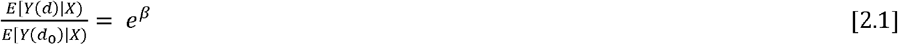

If *X* satisfies the conditional exchangeability assumption, then under model [2.1], *e*^*β*^ also equals the marginal causal risk ratio, allowing valid inference even under selection bias (Robins [14,16]).

We extend the model in Equation [2.1] from two exposure levels to continuous:

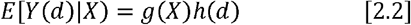

Here, g (X) = *E*[*Y*(*d*_0_)] represents the disease risk at the baseline dose level, and we call it Null Disease Risk (NDR). If *X* satisfies the weak confoundedness assumption, *h*(*d*) can be interpreted as a marginal causal risk ratio function of *d* or a relative causal dose-response curve. We impose the constraint *h*(*d*) = 1, and often assume 0 ≤ *h*(*d*) ≤ 1implying a protective dose effect. A typical choice of *h*(*d*) is a two-parameter logit function 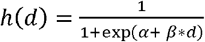. While this motivates the name Covariate Adjusted Logit Model (CALM), the framework can accommodate semi-parametric or non-parametric specifications for both *g*(*x*)and *h*(*t*). Other parametric forms for dose response curves include those in Daniels et al. [1], Cox [17] and Dunning et al. [19].

Although Equation [2.2] provides a causal definition of *h*(*d*), this paper does not pursue causal estimation directly. Instead, we use traditional regression modeling for both *g*(*x*) and *h*(*t*), leave causal development for future research. CALM is thus implemented as following regression model:

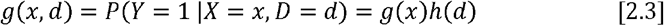

Next, we examine how CALM relates to and other commonly used models and explore its interpretations. If the disease risk does not depend on any covariates, then the NDR *g*(*x*) is a constant and Equation [2.3] simplifies to:

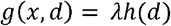

This is known as the Scaled Logit Model (Dunning [18]), used in antibody-disease modeling curve.

In addition to modelinh *h*(*t*) as a function that reduces the NDR *g*(*x*), one can instead define it to reduce the null disease odd 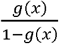. In particular, if *g*(*x*) is modeled via logistic regression, and 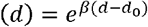, then CALM reduces to a stand logistic regression model, where the interpretation becomes that the null disease odds decline exponentially with increasing dose.

In addition to the conventional interpretation of *h*(*d*) as a function to reduce the null disease risk, vaccine effectiveness research offers an alternative perspective (Halloran et al. [20]; WHO [21], Dunning [18]). Suppose the population consists of two types of individuals: those who are susceptible to disease and those who are immune. In the absence of antibody protection (i.e., at baseline dose), all individuals are assumed susceptible. As the dose level increases, some individuals acquire immunity, while others remain susceptible.

Under this interpretation, *h*(*d*) represents the proportion of individuals who remain susceptible at dose level *d*, and all susceptible individuals with covariate value *x* has a probability *g*(*x*) of developing disease. This framework reflects the so-called All-or-Nothing effect of vaccine protection.

In the All-or-Nothing framework, an individual’s disease risk is governed by their latent binary susceptibility status. If this latent status were observable, estimating *h*(*d*) would no longer require adjustment for confounding covariates. Thus, adjusting for confounders in CALM is equivalent to inferring each individual’s unobserved susceptibility. This conceptualization motivates the Gibbs-style estimation procedure described in the next subsection.

### 2.2. Computation methods

When both components of CALM: *g*(*x*) and *h*(*d*) are specified parametrically, the joint likelihood function is fully tractable, allowing model parameters to be estimated via standard nonlinear maximum likelihood procedures. Bayesian modeling is also feasible under this parametric formulation. However, building on the or-Nothing (AoN) algorithm—to perform Bayesian estimation of *h*(*d*). This approach offers greater flexibility All-or-Nothing interpretation introduced above, we propose a Gibbs-style algorithm—referred to as the All- and is particularly well-suited for accommodating complex or non-parametric structures in the dose-response function.

For observed triplets (*d*_*i*_,*x*_*i*_,*y*_*i*_), *i* = 1,…,*n*, we introduce a latent binary variable *z*_*i*_ to indicate individual *i* is susceptible (*z*_*i*_ = 1) or immune (*z*_*i*_ = 0). Disease can occur only if z =1. We assume that for susceptible individuals, the disease risk is g(x), while the proportion of susceptible individuals at dose level t is *h*(*d*). The conditional probability of *y*_i_ given *x* and *z* is given by:

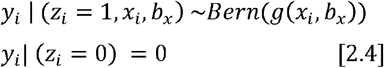

Here, *b*_X_ is the parameter vector of *g*(*x*). The distribution of the susceptible status *z*_*i*_ at dose level *d*_*i*_ is determined by the function *h*(*d*) with parameter *b*_d_:

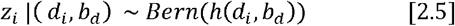

Priors are placed on parameters: *b*_*x*_ ~ *p*(*s*_*x*_), *b*_*d*_ ~ *p*(*s*_*d*_), resulting in the joint likelihood function:

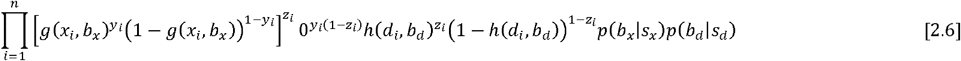

with a notation specification that 0^0^ = 1.

Hence, the posterior distribution of the susceptible status *z*_*i*_ given *d*_*i*_ and the function values of *g*(*x*_*i*_,*b*_*X*_) and *h*(*d*_*i*_,*b*_*d*_):

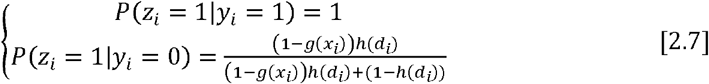

The posterior distribution for *b*_*x*_ is

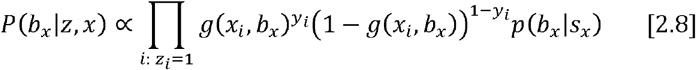

and for *b*_*d*_ :

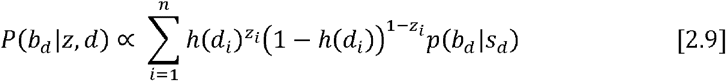

Equations 2.7 to 2.9 defines the AoN Algorithm—a two-step Gibbs sampler cycling between Bayesian regressions on the latent susceptibility indicators. Particularly, the susceptible subjects (*z*_i_ = 1) will be used to update the model parameters for NDR *g*(*x*), and the susceptible status of all subjects is used to update the model parameter of *h*(*d*). If logistic regression models are used, efficient Polya-Gamma sampling algorithm (Polson et al. [22]; Choi and Hobert [23]; Fruhwirth-Schnatter et al. [24]) can be used. For non-parametric estimation of h(d), we can use isotonic regression within the Bayesian framework (Neelon and Dunson [25]; Li and Fu [26]).

## 3. Simulation and GBS Data Analysis

In this section, we begin with simulation experiments to assess whether confounding effects can be appropriately adjusted under selection bias using CALM’s modeling structure. Following this, we apply the CALM model to a real-world data set.

### 3.1. Simulation Study to Evaluate Sampling Bias on CALM Model

To simplify and underscore our main point, we introduce a single binary covariate, *X*, as a confounder. The study population comprises two groups: a high-risk group, *P*(*X*= 1) = 0.25) and a low-risk group, *P*(*X*= 1) = 0.75. The conditional distributions of the dose levels are *f* (*d* |*X* = 1) ~ *N* (−4,1.5) and *f* (*d* |*X* = 0) ~ *N* (0.5,2) respectively. The baseline dose level is set at *d*_0_ = −6.5, and any simulated values below −6.5 are truncated. The top-left panel of Figure 1 illustrates the dose level distributions for the two risk groups.

**Figure 1:**
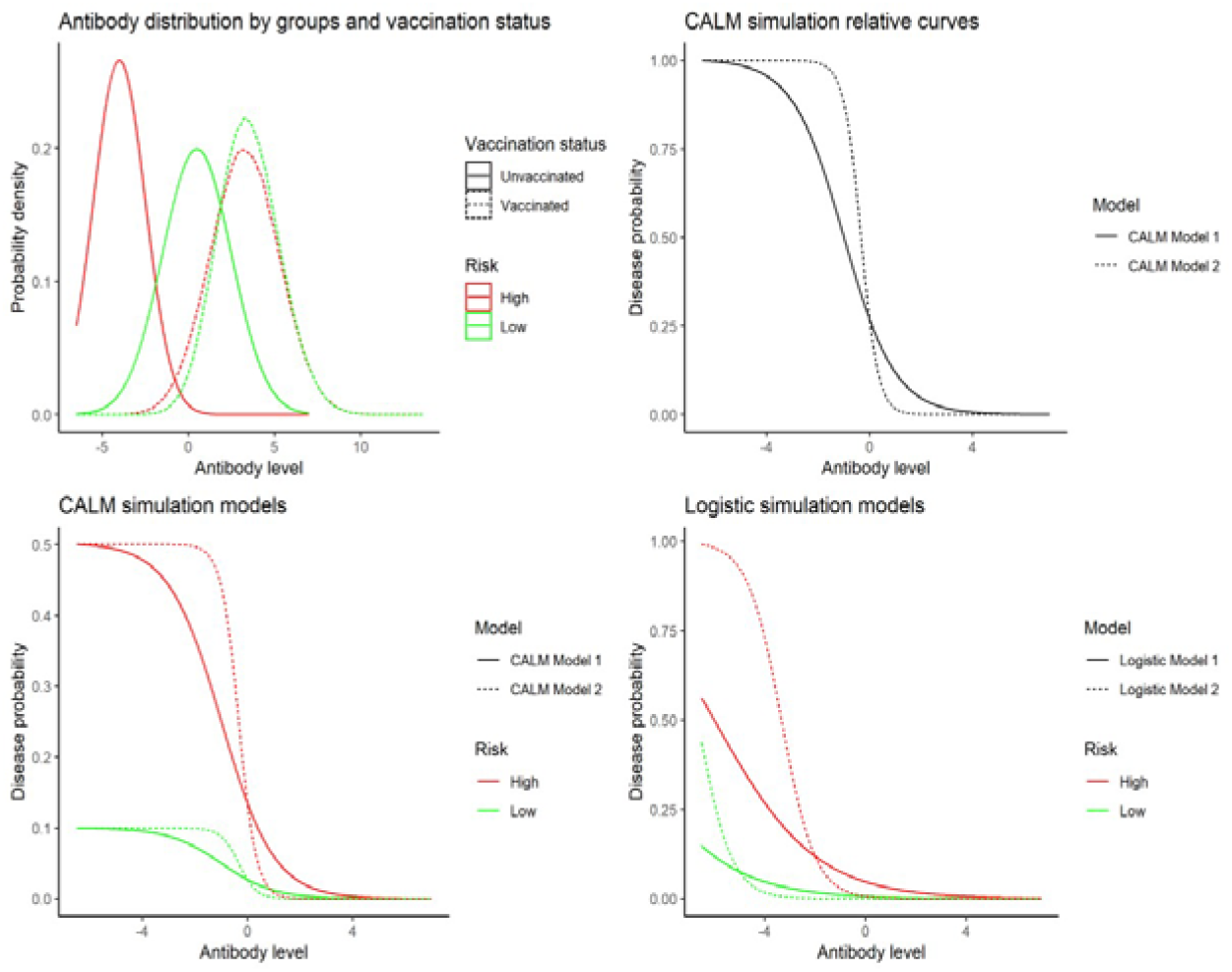
Simulation Design: conditional antibody distributions and disease risk functions

The underlying disease risk follows a CALM model with the null disease risk (NDR) 0.5 for the high-risk group and 0.1 for the low-risk group. The relative dose-response curve is modeled as *h*(*d*) = *h*_0_(*t*)/ *h*_0_ −6.5, and *h*_0_(*d*) = 1/(1 + exp (*α* + *β* * *d*)). Two sets of parameters are considered: Model 1 (*α* = 1, *β*= 1) and Model 2 (*α* = 1, *β*= 3). Figure 1’s top-right panel displays *h*(*d*) for the two models and the bottom-left panel shows the full disease risk function.

To test CALM’s robustness against biased sampling, we simulate observational samples with distorted high-risk proportions: 0.1 and 0.4 instead of the true population value of 0.25. To isolate estimation bias caused by sampling, we simulate two large samples (n = 10,000) for each bias scenario. Random variability from small sample sizes is assessed later.

Four estimation methods are applied to each sample: 1) Parametric CALM with true function form h(t), estimated via MLE; 2) Parametric CALM estimated via Bayesian methods, using non-informative priors (N(0,10^2^)) for the parameters; 3) Non-parametric step function approach using 20 bins (each with 500 subjects), with Beta(1/2,1/2) priors; 4) Non-parametric smoothing spline approach. The AoN algorithm is used for Bayesian estimation in both non-parametric models.

Figure 2 (top panels) shows the estimated *h*(*d*) curves with each biased scenario. All four CALM models approximate the true *h*(*d*) closely, indicating effective adjustment for confounding and sampling bias, even for the two non-parametric methods. The bottom-left panel of Figure 2 presents Bayesian estimates with 95% credible intervals under both biased scenarios. Bias due to sample selection appears negligible.

**Figure 2:**
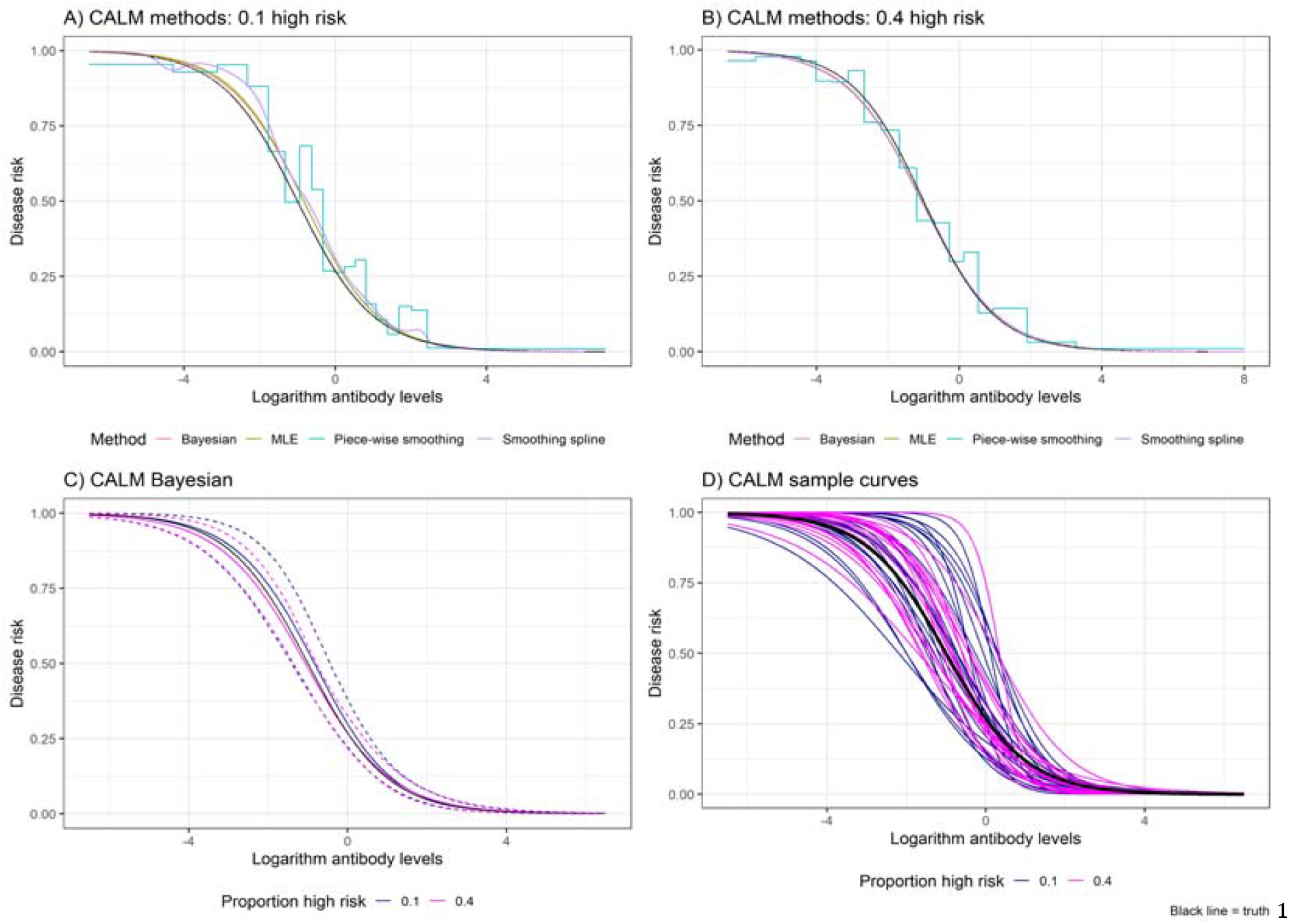
Performance of CALM in adjusting confounding and sample bias ^1^In each graph, the black solid line is the true dose response curve that the simulation data was generated from. A large sample size (n=10,000) is used in panel A,B and C, to focus on CALM’s ability to adjust for sampling bias ^2^A total of 40 estimated curves in panel D – 20 with each sample bias setting. Sample size is 1,000 with approximately 110 observed cases in each sample.

To evaluate smaller-sample performance, we simulate 20 samples of n = 1,000 for each bias scenario. With roughly 110 diseased cases per sample, results from parametric CALM estimation are shown in the bottom-right panel of Figure 2. Despite greater variability due to randomness, estimates still align with the true h(d), and differences across bias levels are small.

### 3.2. Antibody-Disease Risk Curve for US Infant Group B Streptococcus (GBS) study

Our work is motivated by the need to establish antibody-disease risk curves to support maternal vaccine licensure for preventing infant Group B Streptococcus (GBS) disease. Anti-GBS antibodies transferred from vaccinated mothers protect newborns. Our primary data source is a CDC ABCs-based case-control study (Rhodes et al. [9]), with approximately a 1:3 case-to-control ratio. Cases are early-onset GBS cases; controls are healthy infants born to GBS-colonized mothers in selected research centers. The goal is to estimate a protection curve for U.S. infants born to colonized mothers.

Sampling bias exists due to the overrepresentation of some geographic regions (e.g., one state contributes >50% of controls but represents only 5% of the national population). Confounders (gestational age, chorioamnionitis) were selected using a directed acyclic graph (DAG), and geographic region was added to adjust for sample imbalance. All three variables were dichotomized due to small sample size.

We illustrate CALM with interim data for serotype IA (39 cases, 202 controls). Of the 241 subjects, 57 (19 cases, 38 controls) had antibody levels below the detection limit. This subset allows rough stratified estimation of NDR by covariates. Table 1 shows NDR variation by region. Differences in mean log antibody levels by stratum are also included in Table 1.

**Table 1:** Null disease risk and antibody distributions by covariates for GBS study.

| Covariates | strata | Percentage of cases among those with minimal antibody | p-values for percentage of cases between strata of covariates | Mean of logarithm of antibody values | p-values to compare antibody distributions |
| --- | --- | --- | --- | --- | --- |
| Chorioamnionitis | 1 | 0.455 | 0.35 | -2.59 | 0.14 |
|  | 0 | 0.304 |  | -1.54 |  |
| Geographic region | 1 | 0.176 | 0.002 | -1.49 | 0.23 |
|  | 0 | 0.565 |  | -2.09 |  |
| Gestational age | 1 | 0.362 | 0.33 | -1.48 | 0.02 |
|  | 0 | 0.200 |  | -3.22 |  |

<sup>1</sup>The percentages of cases at baseline dose level is presented, as this is a case-control study design.

To assess whether the antibody-disease relationship depends on covariates, we performed isotonic regressions stratified by each covariate. Figure 3’s left panels show unscaled curves; the right panels show curves scaled at the lowest antibody level. Scaled curves closely align, supporting CALM’s assumption that h(t) is invariant to covariates. We further tested this via likelihood ratio tests using interaction terms in CALM. P-values were 0.82 (chorioamnionitis), 0.67 (region), and 0.14 (gestational age), supporting the no-interaction assumption.

**Figure 3:**
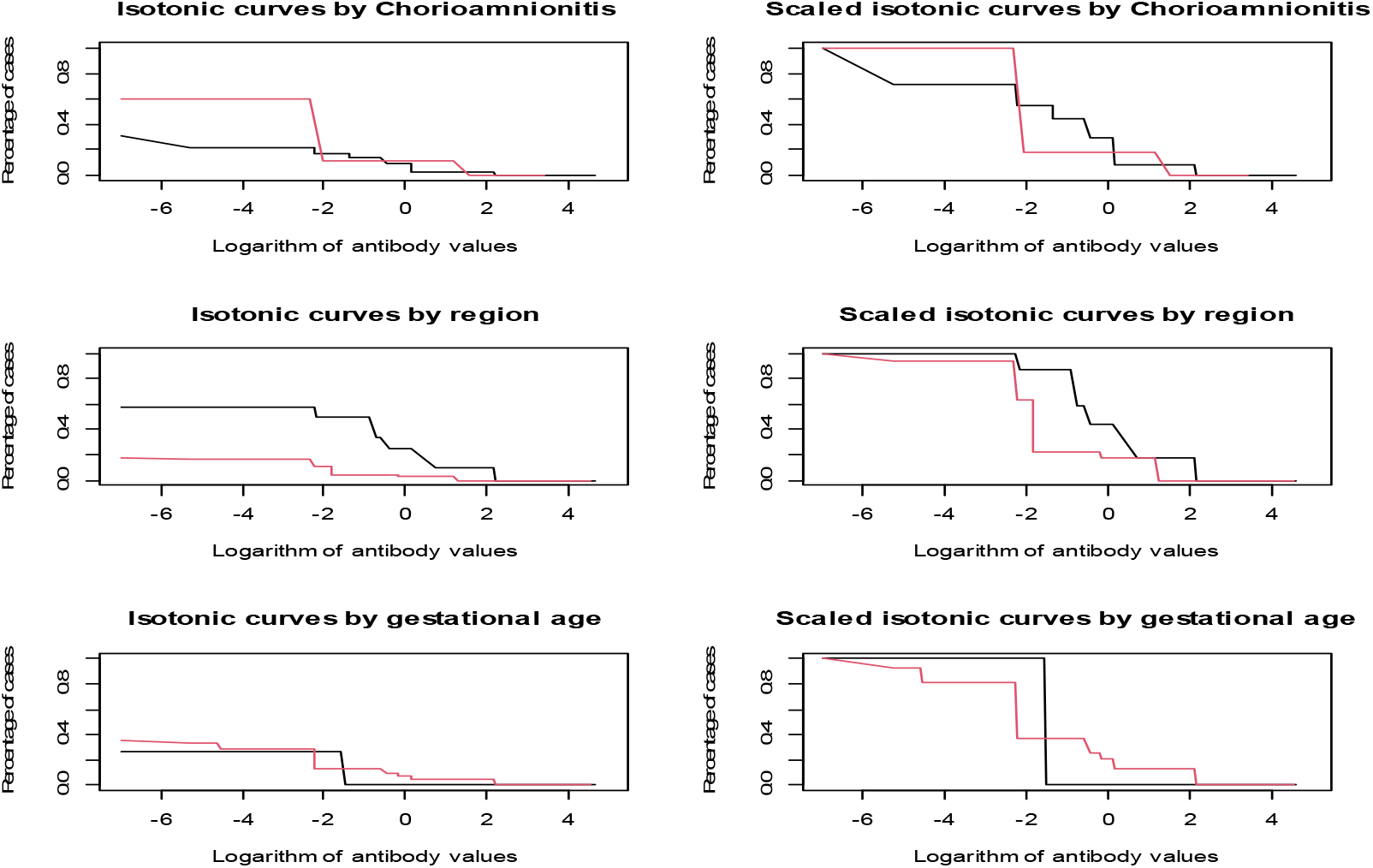
Isotonic regression of antibody levels stratified by covariates for GBS study ^1^The curves in the three right panels are scaled at the lowest antibody level

Figure 4 displays the covariate-adjusted relative dose-response curve (red) estimated via CALM, compared to unadjusted fits from a scaled logit model (black) and isotonic regression (green). The close agreement between the two unadjusted models shows no strong violation in using a logit function to model h(d). The difference between adjusted and unadjusted models highlights the value of covariate adjustment.

**Figure 4:**
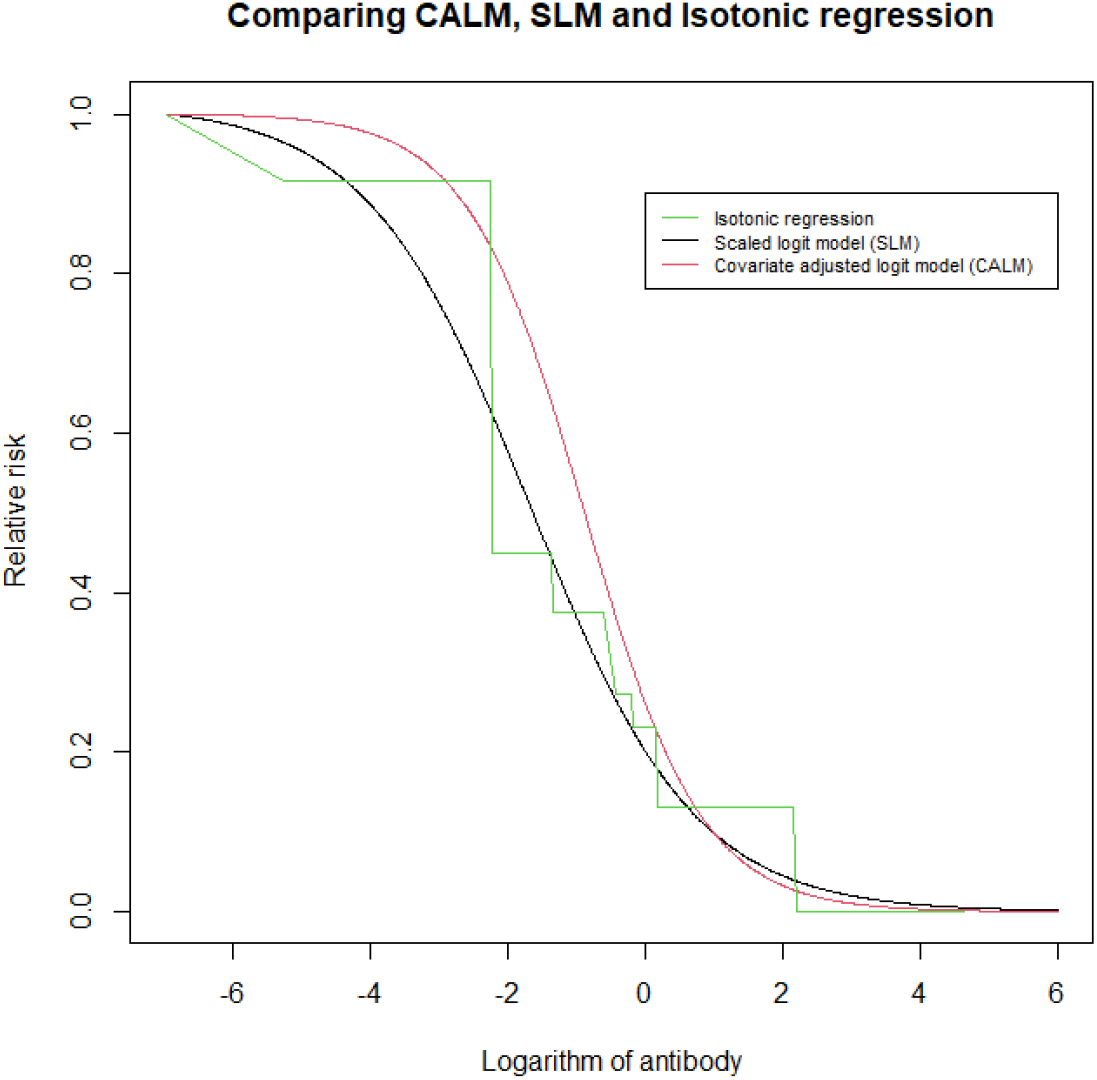
Antibody disease curves constructed by CALM, SLM, and isotonic regression

## 4. Applications of CALM’s Relative Dose-Response Curve in Vaccine Clinical Trials

Traditionally, vaccines are licensed based on demonstrated efficacy in preventing clinical disease endpoints. However, scenarios such as evaluating next-generation vaccines against existing standards, developing vaccines for low-incidence diseases, or responding to biological threats may hinder the ability to demonstrate clinical protection. In such cases, immunologic response can serve as a surrogate for licensure evidence—a concept acknowledged by regulatory bodies (FDA and CBER [27], Rajam et al. [28], Miller et al. [29], Cherry et al. [30]).

Consider a two-arm vaccine trial comparing the vaccine (*v*) and placebo (*p*) arms. Each subject has a disease outcome, an immune response *t*, and a set of covariates *x*. Their joint distribution for arm *a*(*a* = *v* or *p*) is:

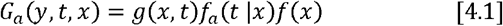

Here, *g*(*x*) = *p*(*y* = 1 | *x,t*)denotes disease risk; *f*_*a*_ (*t* | *x*) is the conditional distribution of antibody levels in arm *a*; and *f* (*x*) is the marginal covariate distribution, identical across arms due to randomization. Vaccine effectiveness (VE) is defined as the reduction of disease risk in the study population:

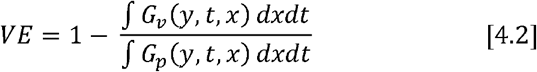

The goal to identify an immunological surrogate is to find an immunological response function *g*(*t*) and use it to estimate the VE as in following equation:

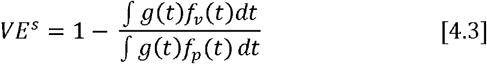

with *f*_*a*_ (*t*) the marginal antibody distribution in arm *a, a* = *v* or *p* such that *VE*^*s*^ = *VE*.

In clinical trial practice, the function *g*(*t*) is further approximated by a binary threshold function such that immune response below a certain level indicates vaccine failure (Qin et. al [31], Siber et al. [32], Chen et al. [33], Donovan et al. [34], Van der Laan et al. [35]).

If there are no confounders—if x does not affect disease risk as: *g*(*x,t*) = *g*(*t*) or *x* does not alter the antibody distribution in both study arms: *f*_*a*_ (*t* | *x*) = *f*_*a*_ (*t*) then existence of a *g*(*t*) such that *VE*^*s*^ in Equation 4.3 is the same as the VE in Equation 4.2 is trivial. For instance, we may use *g*(*t*) = ∫ *g* (*x,t*) f (*x*) *dx*.

However, with the presence of confounders, existing of such function *g*(*t*) remains an unexplored and unanswered q uestion. In a clinical trial, randomization ensures equality in the marginal covariate distribution across study arms, but not in the conditional antibody distribution *f* (*t* | *x*). In fact, covariates unanswered question. In a clinical trial, randomization ensures equality in the marginal covariate may affect vaccine induced immune responses very differently than naturally induced responses. *g*(*t*) can consistently serve as immunologic surrogates under confounding and selection bias. To address this, Despite the importance of the issue, there has been little mathematical validation of whether such a function *g*(*t*) can consistently serve as immunologic surrogates under confounding and selection bias. To address this, we extend the simulations from Section 3 to evaluate multiple surrogate construction methods under known VE settings.

### 4.1. Overview of surrogate construction approaches

We focus on constructing *g*(*t*) using observational data from the unvaccinated population, as in many practical cases (e.g., GBS studies). Assuming no unmeasured confounders and conditional exchangeability, we derive *g*(*t*) from the estimated risk function *g*(*t,x*). Ten approaches are evaluated and grouped into four categories:

Group 1 (Gold Standard): Use the estimated joint risk function *ĝ*(*x,t*) to estimate VE with Equation 4.2. It is considered gold standard, as the true function form that the data was simulated from will be used to model the observed data.

Group 2 (Ignore Covariates): *g*(*t*) will be estimated directly ignoring the covariates. Two regression methods are considered: logistic regression and scaled logit model g(t) = λh(t).

Group 3 (Logistic Regression + Marginalization): After applying a logistic regression model logit (g(x,t)) = *α* + *β* * *t γ* * *x* to obtain *ĝ*(*x,t*), following four approaches will be used to derive *ĝ*(*t*): a) Drop the covariate term from the estimated function: 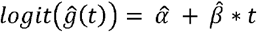. Under our simulation design, this is equivalent to use the covariate-specific curve in the low-risk group (*x* = 0)b) Average intercept with sample marginal distribution: 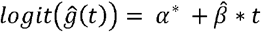 where 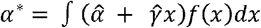, where *f* (*x*) is the marginal distribution of *x* in the sample; c) Apply marginalization g-formula: *ĝ*(*x,t*) = ∫ *ĝ*(*x,t*) *f*_*p*_ (*t* | *x*) *dx*, using the conditional antibody distribution among unvaccinated people. Notice that this *ĝ*(*t*) is derived such that the denominators in Equation 4.2 and Equation 4.3 are equal; d) Use separate *ĝ*(*t*) for unvaccinated and vaccinated groups. For unvaccinated group, 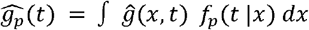 and in the vaccinated group, we assume vaccine-induced antibody distribution does not associated with the covariate, hence 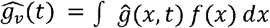.

Group 4 (CALM-Based): Here *ĝ*(*x,t*) is estimated under the CALM modeling structure: *ĝ*(*x,t*) = *ĝ*(*t*) *h* (*t*). Then *ĥ* (*t*) will be used as *g*(*t*) in Equation 4.3. We also apply the two g-formula approaches in Group 3 (approach c and d) to CALM’s estimated *ĝ*(*x,t*).

### 4.2. Simulation to compare immunologic surrogate approaches for VE estimation

We extend Section 3’s simulation to introduce a vaccine with known VE = 80%. For unvaccinated individuals, antibody distributions are identical to before. For vaccinated individuals, the vaccine induced antibody levels follow *N* (3.5,2). Their observed antibody levels are set as the maximum of vaccine- and naturally induced levels. The antibody distributions are presented in Figure 1 (top-left panel). The distributions are less prone to the covariate among vaccinated subjects.

The underlying disease risk function *g*(*x,t*) is simulated under the two CALM models as presented in Section 3 as well as under two additional logistic regression model *logit* (*g*(*x,t*)) = *α* + *β t* + *γ x*, with parameter combinations: Model 1: α = −1, β = −0.5, γ = −2 and Model 2: α = 0, β = −1.5, γ = −5. The four models are illustrated in the lower-right panel of Figure 1.

We adopt a similar sampling scheme as in Section 3, generating samples of size 1,000, each containing approximately 110 disease cases. For each of the four risk models, we simulate two scenarios with different marginal covariate distributions: an unbiased case with *P* (*X* = 1) = 0.25 and a biased case with P(X=1)=0.10. This yields a total of 8,000 simulated datasets (1,000 repetitions per model and bias scenario). For each dataset, the ten surrogate construction approaches described in Section 4 are applied to derive immunologic surrogate functions. These surrogates are then used to estimate vaccine effectiveness (VE) in a hypothetical target population of 200,000 individuals, with 50% randomly assigned to vaccination.

Figure 5 presents vaccine effectiveness (VE) estimates under unbiased sampling conditions, with each panel corresponding to one of the four simulation models. Methods in Group 2, which ignore covariates, exhibit systematic bias—highlighting that covariate adjustment is essential. Group 3, which uses logistic regression models with various marginalization techniques, also performs poorly, even when the model is correctly specified (as seen in the lower two panels). Notably, these results reveal a key limitation of applying the g-formula to the absolute disease risk function: deriving a common function g(t) from unvaccinated subjects and applying it to both treatment arms can be problematic. When confounding covariates affect the antibody distribution differently across vaccine arms, the assumption of a single absolute risk function becomes untenable, and bias is introduced by extrapolating from unvaccinated data alone.

**Figure 5:**
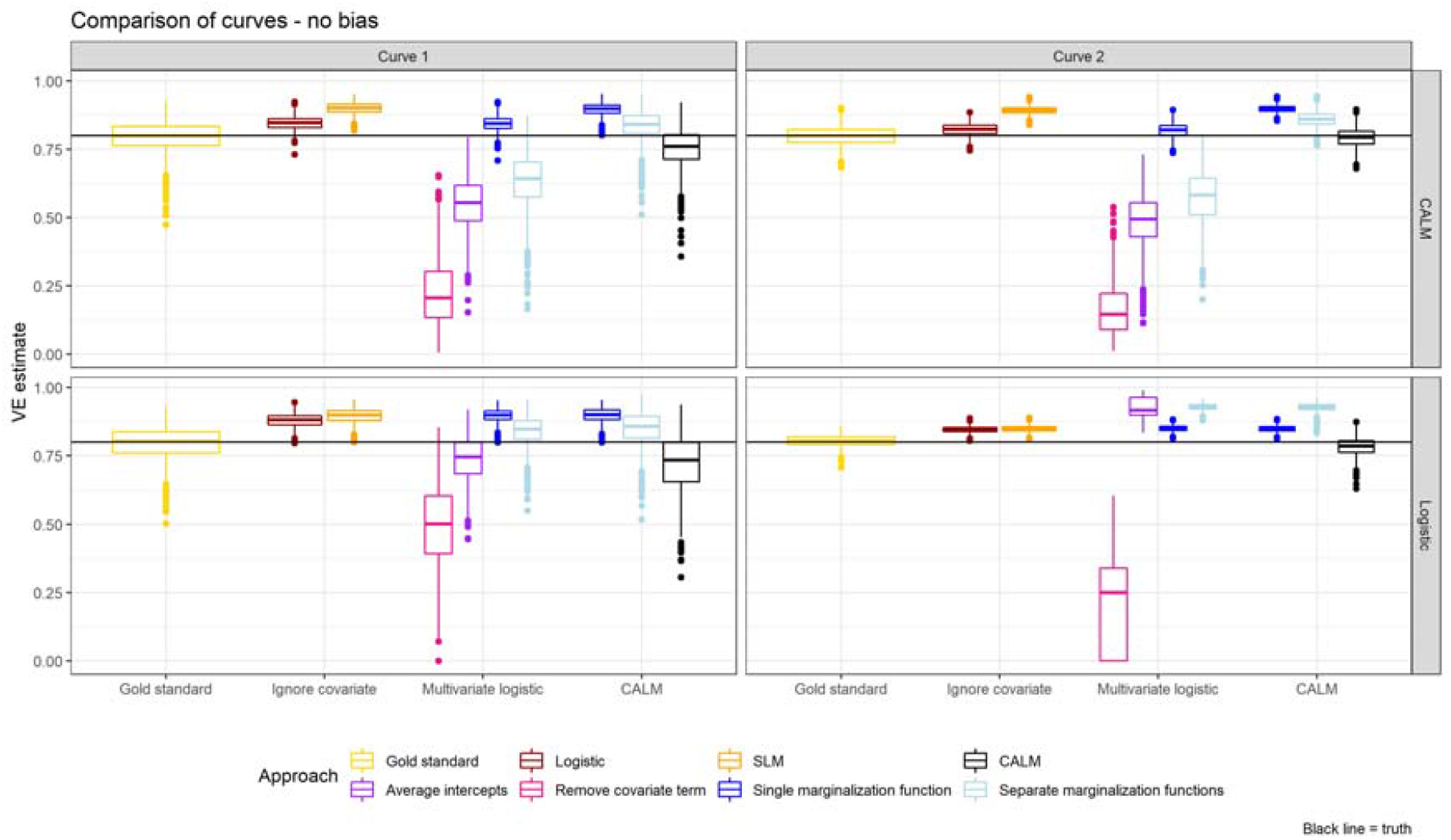
VE Estimations by different approaches with unbiased sample ^1^The four panels correspond to the four underlying simulation models: the top two panels follow the CALM structure, while the bottom two follow standard logistic regression model. ^2^The gold standard is the full join disease risk function g(x, t), estimated using the same function forms as those used to generate the simulated data. ^3^ The single marginalization function approach applies the same marginalized function to both vaccinated and unvaccinated subjects. The function is derived solely from observational data of unvaccinated subjects. ^4^The separate marginalization function approach uses different marginalized functions for vaccinated and unvaccinated subjects. For unvaccinated subjects, the function is chosen to ensure that the denominators in Equation 4.2 and 4.3 are equal. For the vaccinated subjects, antibody levels are assumed to be independent of covariates; thus, the marginalized function does not need observations from the vaccinated subjects, as the marginal distribution of covariates are equal across the study arms through randomization. ^5^ The horizontal black line indicates true VE of 80%

In contrast, the CALM-based h(t) curves consistently outperform other approaches, even when the underlying simulation model does not conform to the CALM structure. This robustness highlights the advantage of using a relative dose-response framework for estimating vaccine effectiveness (VE), which is inherently a relative measure. Moreover, CALM offers the additional benefit of effectively adjusting for both confounding and sample selection bias.

Figure 6 compares results under biased versus unbiased sampling. Marginalization-based approaches show sensitivity to sampling bias. CALM-based relative h(t) is more robust.

**Figure 6:**
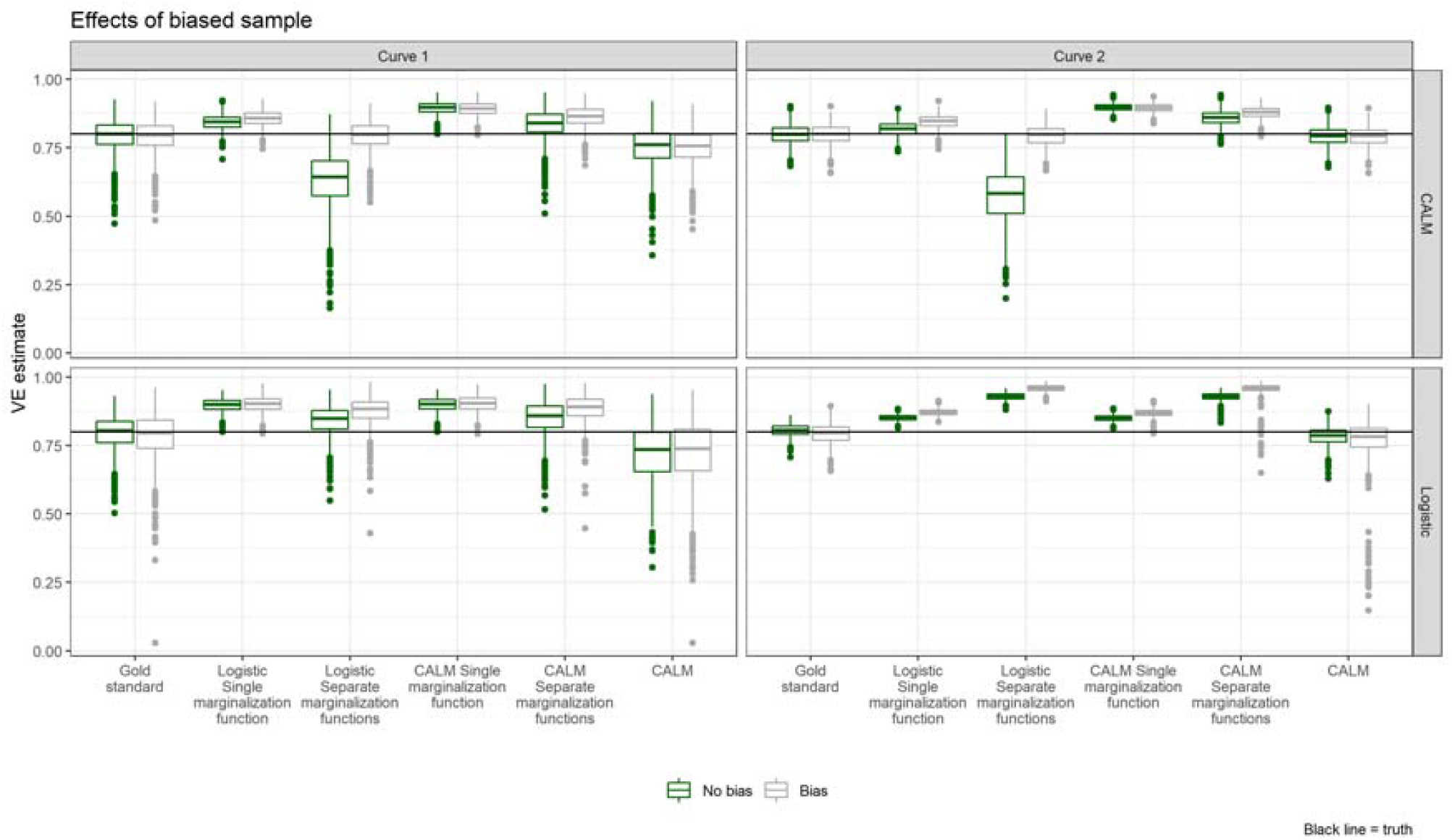
Comparing VE estimation approaches with biased and unbiased sample

Table 2 presents MSEs of VE estimates when compared to the gold standard. CALM-based curves show the lowest errors, validating their reliability for surrogate construction under confounding and selection bias.

**Table 2:** Mean Square Errors of VE Estimations.

| Methods | CALM |  |  |  | Logistic |  |  |  |
| --- | --- | --- | --- | --- | --- | --- | --- | --- |
|  | Model 1 |  | Model 2 |  | Model 1 |  | Model 2 |  |
|  | unbiased | biased | unbiased | biased | unbiased | biased | unbiased | biased |
| Logistic, no covariate | 0.005 | 0.01 | 0.002 | 0.005 | 0.009 | 0.019 | 0.002 | 0.007 |
| Scaled logit model | 0.014 | 0.013 | 0.01 | 0.009 | 0.013 | 0.02 | 0.002 | 0.007 |
| Logistic, remove covariate term | 0.331 | 0.199 | 0.409 | 0.192 | 0.103 | 0.099 | 0.394 | 0.368 |
| Logistic, marginalized intercept | 0.073 | 0.031 | 0.107 | 0.025 | 0.013 | 0.024 | 0.016 | 0.018 |
| Logistic, single marginalization function | 0.005 | 0.007 | 0.002 | 0.003 | 0.013 | 0.019 | 0.002 | 0.007 |
| Logistic, two marginalization function | 0.032 | 0.001 | 0.06 | 0.001 | 0.002 | 0.011 | 0.015 | 0.028 |
| CALM, single marginalization function | 0.013 | 0.013 | 0.011 | 0.01 | 0.014 | 0.02 | 0.002 | 0.007 |
| CALM, two marginalization functions | 0.002 | 0.006 | 0.004 | 0.007 | 0.005 | 0.014 | 0.015 | 0.028 |
| CALM, relative dose response curve | 0.002 | 0.002 | 0 | 0 | 0.01 | 0.006 | 0.001 | 0.008 |

<sup>1</sup> The mean square errors are calculated between the estimated VEs and the VEs obtained from gold standard. The gold standard is the full join disease risk function $g(x, t)$ , estimated using the same function forms as those used to generate the simulated data

## 5. Discussion

Establishing dose-response relationships for causal relationships using observational data involves addressing two major challenges: confounding and sample selection bias. When selection bias leads to discrepancies in the marginal distribution of confounding covariates between the observational sample and the target population, standard causal inference approaches -- such as g-formula marginalization and propensity scores – may yield biased estimates. To address this, we propose a modeling framework following a general practice in medical and epidemiological research that using logistic regression model to obtain “adjusted” odds ratio.

The first component of our strategy is the use of a relative dose-response curve, which expresses disease risk at each dose level relative to a baseline. While this approach does not yield absolute risk estimates, it preserves the essential shape and interpretability of the exposure-outcome relationship. Importantly, relative risk measures—such as risk ratios and odds ratios—are standard in epidemiology, making this a natural and intuitive framework.

The second component is a more restricted definition of confounding. In CALM, a covariate is considered as a confounder only if it is associated with both the exposure and outcome and does not interact with the relative dose-response curve. If the relative curve varies substantially across covariate values, then constructing stratified dose-response curves is preferable to marginalizing over a potentially non-representative sample.

These two components motivate the Covariate-Adjusted Logit Model (CALM), which explicitly separates the null disease risk (NDR) from the relative response curve. This framework generalized the log-linear structural mean model for binary exposures to continuous exposures and ensures that the marginal and conditional relative risk functions coincide under the weak exchangeability assumption. Because the relative response function is invariant to the marginal distribution of covariates, CALM enables valid estimation even in the presence of sample selection bias – a property confirmed through our simulation studies. The model’s no-interaction assumption can be assessed visually e using stratified non-parametric analyses (e.g., isotonic regression) or tested via likelihood ratio tests through including interaction terms into CALM modes.

Beyond confounding and sample selection bias, absolute dose-response modeling faces an additional challenge when attempting to apply a common curve across different populations with differing exposure-covariate associations. This situation arises in vaccine clinical trials when immunological surrogates are used to estimate vaccine effectiveness. While randomization balances marginal covariate distributions, it does not ensure comparable conditional antibody distributions – especially if vaccine-induced immune response is influenced by covariates differently than naturally induced response. In such settings, no shared absolute dose-response curve may exist to support unbiased VE estimation.

In contrast, CALM facilitates the construction of a shared relative response curve that reflects a meaningful biological quantity: the reduction in latent susceptibility. This interpretation aligns directly with VE, which is itself a relative quantity, making CALM especially useful in future vaccine trials using immunological surrogates. Our simulations results demonstrate that CALM consistently outperforms alternative approaches in the presence of confounding and sample selection bias.

This interpretation of the relative dose-response curve as a function that captures reductions in latent susceptibility not only motivates the development of a Gibbs-style algorithm (All-or-Nothing algorithm) for Bayesian implementation of CALM, but also offers conceptual insights for future research on confounding adjustment within the casual inference framework. Specifically, under CALM, adjusting for confounding can be interpreted as an attempt to uncover each individual’s latent susceptibility status. We plan to further explore this idea in future work.

In conclusion, CALM offers a flexible, robust, and interpretable framework for estimating dose-response relationships from observational data. By focusing on relative effects and limiting confounding to exclude interactive covariates, CALM provides valid inference when standard approaches fail. Its performance in both real-world and simulation datasets supports its utility in epidemiologic studies and in vaccine evaluation.

## Data Availability

The code to simulate data is available upon request

## Disclaimer

The findings and conclusions in this report are those of the author(s) and do not necessarily represent the official position of the Centers for Disease Control and Prevention.

